# AI-MET: A Deep Learning-based Clinical Decision Support System for Distinguishing Multisystem Inflammatory Syndrome in Children from Endemic Typhus

**DOI:** 10.1101/2023.06.15.23291453

**Authors:** Abraham Bautista-Castillo, Angela Chun, Tiphanie P. Vogel, Ioannis A. Kakadiaris

## Abstract

The COVID-19 pandemic brought several diagnostic challenges, including the post-infectious sequelae multisystem inflammatory syndrome in children (MIS-C). Some of the clinical features of this syndrome can be found in other pathologies such as Kawasaki disease, toxic shock syndrome, and endemic typhus. Endemic typhus, or murine typhus, is an acute infection treated much differently than MIS-C, so early detection is crucial to a favorable prognosis for patients with these disorders. Clinical Decision Support Systems (CDSS) are computer systems designed to support the decision-making of medical teams about their patients and intended to improve uprising clinical challenges in healthcare. In this article, we present a CDSS to distinguish between MIS-C and typhus that includes a scoring system that allows the timely distinction of both pathologies only using clinical and laboratory features typically available within the first six hours of presentation to the Emergency Department (ED). The proposed approach was trained and tested on datasets of 87 typhus patients and 133 MIS-C patients. A comparison was made against five well-known statistical and machine-learning models. A second dataset with 111 MIS-C patients was used to verify the AI-MET effectiveness and robustness. The performance assessment for AI-MET and the five statistical and machine learning models was done by computing Sensitivity, Specificity, Accuracy, and Precision. The AI-MET system scores 100 percent in the five metrics used on the training and testing dataset and 99 percent on the validation dataset.

## 1 Introduction

In December 2019, in Wuhan, China, the first Coronavirus disease 2019 (COVID-19) cases were reported [1], and several clinical challenges subsequently appeared. At that time, even though it was recognized that children were as susceptible to COVID-19 as adults, this infection was less frequently observed in pediatric patients than in adults (1-5% of all COVID-19 cases) [2], and tended to present with a milder clinical course [3]–[6].

However, in April 2020, some children began to be hospitalized at an increased rate due to fever and multisystem inflammation [7]–[10], and in May 2020, the Centers for Disease Control and Prevention (CDC) published a case definition for multisystem inflammatory syndrome in children (MIS-C) [11].

Some of the most frequent features found in MIS-C patients are fever, rash, conjunctivitis, oromucosal changes, abdominal pain, vomiting, diarrhea, myocarditis, and hematologic abnormalities [7]–[12]. These findings overlap other diseases such as Kawasaki Disease (KD) [13], toxic shock syndrome (TSS) [14], and murine typhus [15], [16], provoking a clinical challenge to distinguish MIS-C from these pathologies.

Clinical Decision Support Systems (CDSS) are computer systems designed to support the decision-making of medical teams about their patients and intended to improve care with clinical knowledge and patient information [17]. These systems are often divided into knowledge and non-knowledge systems, where knowledge systems are those that make use of information provided by medical personnel to create the rules they use. In contrast, non-knowledge systems use artificial intelligence, machine learning, or statistical methods *in lieu* of instructions obtained from medical personnel [18], [19]. Advances in these fields have significantly increased the presence of CDSS in the medical field [20], to address clinical challenges such as antibiotic management [21], heart disease prediction [22], and cancer detection [23].

With its clinical and laboratory similarities to MIS-C, distinguishing typhus has become a diagnostic challenge in the US where it is endemic: Texas, southern California, and Hawaii [24]. Timing has become crucial, as early identification and targeted therapy are essential for better outcomes in both conditions [25], [26]. This opens the opportunity for the implementation of a CDSS that can help the medical staff in the ED with timely decision-making.

The main contributions of this paper are:

1. Developing a novel two-stage CDSS for distinguishing between MIS-C and typhus.
2. Creating a fixed-score system capable of being implemented in the ED without an electronic device.
3. Optimizing the number of clinical features required to differentiate MIS-C from typhus.

## 2 Related Work

CDSS has been used during the COVID-19 pandemic as support tools for the prognosis of disease severity [27] for predicting mortality [28]. However, only a few have focused on MIS-C-related clinical challenges.

Soneji *et al*. [29] used Random Forest as a model to achieve a high sensitivity MIS-C diagnosis and to provide insights about the importance of features such as procalcitonin, ferritin, N-terminal pro-B-type natriuretic peptide (NT-proBNP), and C-reactive protein (CRP). A point to consider for CDSS is how missing values are treated, with the median value based on the presence or absence of MIS-C in this approach.

On the other hand, Clark *et al*. [30] proposed to use logistic regression with bootstrap backward selection to identify the most relevant predictors for MIS-C. This model identified patients with MIS-C among an otherwise undifferentiated group of febrile patients with features obtained within the first 24 hours of hospital presentation.

Finally, Lam *et al*. [31] built a two-stage model of feedforward neural networks. This model is intended to differentiate between MIS-C, KD, and children with nonspecific febrile illnesses. In the first stage of this model, the patient can be classified as MIS-C, not MIS-C, or rejected by the conformal prediction framework. If the patient is classified as not MIS-C, data will pass to the next stage and be used to differentiate between a febrile child and a KD patient. For the conformal prediction framework, they used trust sets by filtering patients in the training cohort with more than one missing value and an MIS-C risk score greater than the 95^*th*^ percentile.

Even though these CDSS focused on the clinical challenge that MIS-C represents, none considered endemic typhus as one of the possible overlapping febrile diseases of childhood, and none presented an alternative tool to be implemented in the ED without using electronic devices. Only Clark *et al*. considered the importance of timely prediction using features obtained within the first 24 hours. Therefore, a CDSS capable of distinguishing between patients with MIS-C or typhus during the first hours of their presentation and implementation without the need for an electronic device is necessary.

## 3 Methods

AI-MET is a decision support system divided into two main stages, MET-17 and MET-30 (Fig. 1). During the first stage, a fixed scoring system provides patients’ classification and a confidence index for this classification without using a computer system or electronic device. This stage is named MET-17. If the classification doesn’t meet the established confidence index, the medical team can appeal to the next stage, MET-30. MET-30 is a recurrent neural network with an attention module capable of distinguishing patients with MIS-C from patients with typhus and computing the importance of its inputs for this classification. Next, we present the pre-processing, feature selection, and methods for AI-MET.

**Figure 1:**
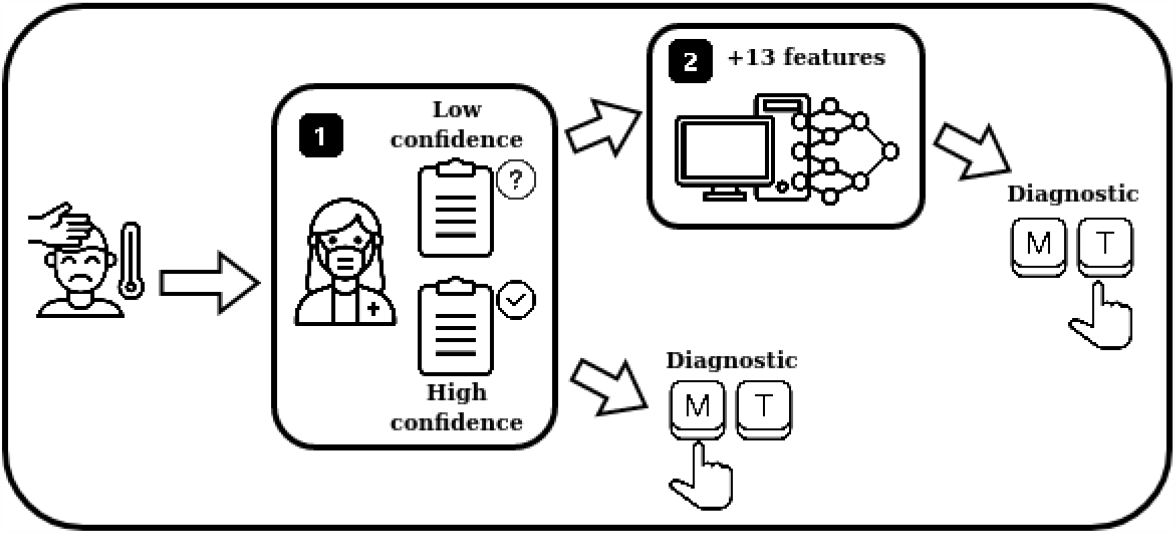
Flowchart of the two-stage decision support system AI-MET.

### 3.1 Pre-processing

The pre-processing performed on the dataset comprises the categorization and normalization of the patient features. For the feature categorization process, we used predefined thresholds that convert a feature into up to three inputs depending on the thresholds used (Fig. 2). Laboratory features such as white blood cells (WBC), absolute lymphocyte count (ALC), absolute neutrophil count (ANC), platelet count, sodium, troponin, B-type natriuretic peptide (BNP), and fibrinogen was categorized as low, normal, and high using the thresholds established in Table 1. In cases such as aspartate aminotransaminase (AST), lactate dehydrogenase (LDH), and alanine aminotransaminase (ALT), we used Tables 2, 3, and 4, which take into account the age and sex to define low, normal, and high. All established thresholds were obtained from the Texas Children’s Hospital (TCH) Pathology Catalog standard references. Demographics and clinical features were normalized using the function “normalize” from scikit-learn package preprocessing using *ℓ*^2^-norm. For categorical features, we established “1” as presence and “0” as absence. Demographic, clinical, and laboratory features provided 73 inputs that were reduced to 40 (see subsection III.D) for the models shown in this study.

**Table 1:** TCH established thresholds to categorize lab test results.

| Laboratory feature | Low | High |
| --- | --- | --- |
| WBC (K/μL) | <4 | >11 |
| ALC (K/μL) | <1500 | n/a |
| ANC (K/μL) | n/a | >8000 |
| Platelets (x1000/μL) | <200 | >500 |
| Sodium (mmol/L) | <135 | n/a |
| Troponin (ng/mL) | n/a | >0.03 |
| BNP (pg/mL) | n/a | >200 |
| NT-proBNP (pg/mL) | n/a | >125 |
| D-dimer (mg/L) | n/a | >0.40 |
| Fibrinogen (mg/L) | <220 | >440 |

**Table 2:**
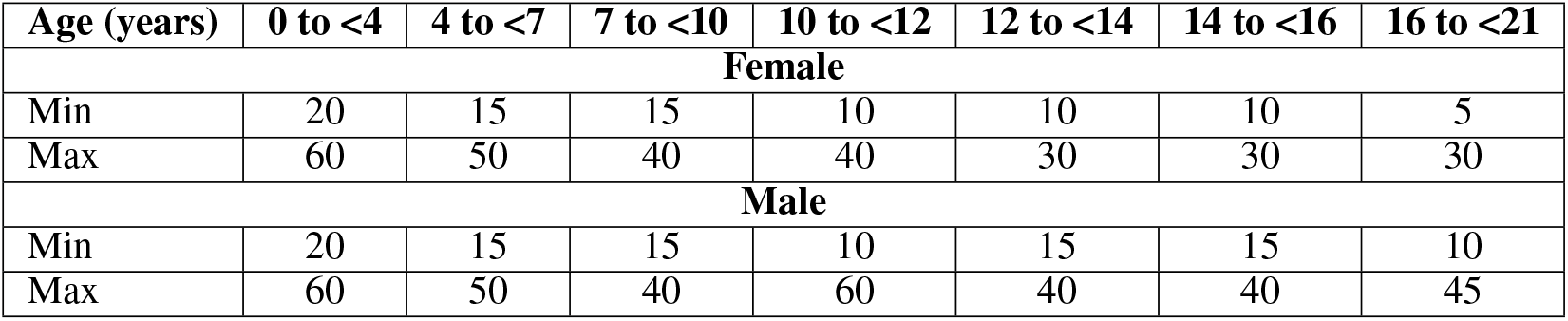
TCH established thresholds to categorize AST (U/L) lab test.

| Age (years) | 0 to <4 | 4 to <7 | 7 to <10 | 10 to <12 | 12 to <14 | 14 to <16 | 16 to <21 |
| --- | --- | --- | --- | --- | --- | --- | --- |
| **Female** |  |  |  |  |  |  |  |
| Min | 20 | 15 | 15 | 10 | 10 | 10 | 5 |
| Max | 60 | 50 | 40 | 40 | 30 | 30 | 30 |
| **Male** |  |  |  |  |  |  |  |
| Min | 20 | 15 | 15 | 10 | 15 | 15 | 10 |
| Max | 60 | 50 | 40 | 60 | 40 | 40 | 45 |

**Table 3:** TCH established thresholds to categorize LDH (U/L) lab test.

| Age (years) | 0 to <4 | 4 to <7 | 7 to <10 | 10 to <12 | 12 to <14 | 14 to <16 | 16 to <21 |
| --- | --- | --- | --- | --- | --- | --- | --- |
| **Female** |  |  |  |  |  |  |  |
| Min | 223 | 209 | 187 | 169 | 169 | 174 | 151 |
| Max | 409 | 401 | 334 | 343 | 285 | 258 | 298 |
| **Male** |  |  |  |  |  |  |  |
| Min | 223 | 209 | 187 | 192 | 209 | 160 | 151 |
| Max | 409 | 401 | 334 | 312 | 334 | 325 | 298 |

**Table 4:** TCH established thresholds to categorize ALT (U/L) lab test.

| Age (years) | 0 to <4 | 4 to <7 | 7 to <10 | 10 to <12 | 12 to <14 | 14 to <16 | 16 to <21 |
| --- | --- | --- | --- | --- | --- | --- | --- |
| **Female** |  |  |  |  |  |  |  |
| Min | 14 | 11 | 11 | 11 | 11 | 10 | 10 |
| Max | 45 | 28 | 28 | 28 | 28 | 35 | 35 |
| **Male** |  |  |  |  |  |  |  |
| Min | 12 | 10 | 10 | 10 | 10 | 11 | 11 |
| Max | 45 | 41 | 41 | 41 | 41 | 26 | 26 |

**Table 5:** Training/Testing Dataset

| ID | Feature | Description | Data Type | Min | Max | MIS-C group (patients = 133) |  |  | Typhus group (patients = 87) |  |  |
| --- | --- | --- | --- | --- | --- | --- | --- | --- | --- | --- | --- |
|  |  |  |  |  |  | Median | IQR | Yes (%) | Median | IQR | Yes (%) |
| 1 | Age (years) | Patient's age when admitted | N | 0.75 | 19 | 9 | 5-12 | - | 12 | 9-15 | - |
| 2 | Sex* | Female = 0, Male = 1 | B | 0 | 1 | Male: 77 (58%)<br>Female: 56 (42%) |  |  | Male: 43 (49%)<br>Female: 44 (51%) |  |  |
| 3 | Epidemiological link to SARS-CoV-2 | Self-reported epidemiological link to a confirmed SARS-CoV-2 case in weeks prior to admission | B | 0 | 1 | - | - | 91 (68%) | - | - | 15 (17%) |
| 4 | Fever days before hospital (days) | Self-reported days of fever before admission | N | 0 | 15 | 5 | 4-6 | - | 7 | 6-9 | - |
| 5 | Fever Tmax (oF) | Highest temperature reported by patient or recorded within two hours of ED arrival | N | 100 | 106.2 | 103.5 | 103-104 | - | 103 | 102-103 | - |
| 6 | Lowest systolic BP in ED (mmHg) | Lowest systolic blood pressure reported within two hours of ED arrival | N | 60 | 134 | 96 | 83-107 | - | 108 | 99-116 | - |
| 7 | Lowest diastolic BP in ED (mmHg) | Lowest diastolic blood pressure reported within two hours of ED arrival | N | 17 | 123 | 53 | 43-59 | - | 58 | 54-66 | - |
| 8 | Highest heart rate in ED (bpm) | Highest heart rate reported within two hours of ED arrival | N | 61 | 200 | 135 | 120-147 | - | 113 | 103-127 | - |
| 9 | Antecedent illness | Self-reported illness in weeks prior to admission | B | 0 | 1 | - | - | 51 (38%) | - | - | 6 (7%) |
| 10 | Conjunctivitis | Redness of the conjunctiva | B | 0 | 1 | - | - | 96 (72%) | - | - | 34 (39%) |
| 11 | Rash | Abnormal change in skin color | B | 0 | 1 | - | - | 68 (51%) | - | - | 70 (80%) |
| 12 | Adenopathy | Enlarged cervical lymph nodes | B | 0 | 1 | - | - | 9 (7%) | - | - | 12 (14%) |
| 13 | Mucosal changes | Strawberry tongue, cracked lips, erythema | B | 0 | 1 | - | - | 38 (28%) | - | - | 13 (15%) |
| 14 | Hand/foot changes | Swelling or erythema of hand/foot | B | 0 | 1 | - | - | 13 (10%) | - | - | 0 (0%) |
| 15 | GI involvement | Nausea/vomiting, abdominal pain, diarrhea | B | 0 | 1 | - | - | 123 (92%) | - | - | 65 (75%) |
| 16 | Neuro involvement | AMS, headache, encephalopathy | B | 0 | 1 | - | - | 75 (56%) | - | - | 58 (67%) |
| 17 | MSK involvement | Arthralgias, arthritis, myalgias | B | 0 | 1 | - | - | 49 (37%) | - | - | 51 (59%) |
| 18 | Respiratory involvement | Shortness of breath, respiratory distress, cough, congestion | B | 0 | 1 | - | - | 47 (35%) | - | - | 33 (38%) |
| 19 | Cardiac involvement | Chest pain, palpitations | B | 0 | 1 | - | - | 14 (11%) | - | - | 8 (9%) |
| 20 | White Blood Cells (WBC) (K/μL) | WBC laboratory test within six hours of presentation | N | 2.77 | 28.3 | 9.41 | 7.18-12.42 | - | 7.3 | 5.8-9.4 | - |
| 21 | Absolute Lymphocyte Count (ALC) (K/μL) | ALC laboratory test within six hours of presentation | N | 0.07 | 15.11 | 0.96 | 0.66-1.66 | - | 1.92 | 1.12-2.72 | - |
| 22 | Absolute Neutrophil Count (ANC) (K/μL) | ANC laboratory test within six hours of presentation | N | 0.38 | 25.02 | 7.39 | 5.02-10.3 | - | 4.72 | 3.40-6.23 | - |
| 23 | Platelets (x1000/μL) | Platelets laboratory test within six hours of presentation | N | 38 | 489 | 155 | 117-206 | - | 161 | 120-224 | - |
| 24 | Aspartate Aminotransaminase (AST) (U/L) | AST laboratory test within six hours of presentation | N | 18 | 7,500 | 52 | 38-77 | - | 121 | 78-192 | - |
| 25 | Alanine Aminotransaminase (ALT) (U/L) | ALT laboratory test within six hours of presentation | N | 9 | 2,951 | 37 | 22-56 | - | 80 | 53-137 | - |
| 26 | Sodium (mmol/L) | Sodium laboratory test within six hours of presentation | N | 120 | 144 | 133 | 130-135 | - | 135 | 133-137 | - |
| 27 | Troponin (ng/mL) | Troponin laboratory test within six hours of presentation | N | 0.01 | 7.44 | 0.05 | 0.01-0.201 | - | 0.01 | 0.01-0.014 | - |
| 28 | B-type Natriuretic Peptide (BNP) (pg/mL) | BNP laboratory test within six hours of presentation | N | 10 | 4,500 | 167 | 41-560 | - | 11.95 | 10-26.2 | - |
| 29 | Fibrinogen (mg/dL) | Fibrinogen laboratory test within six hours of presentation | N | 162 | 953 | 563 | 490-646 | - | 394 | 322-448 | - |
| 30 | Lactate Dehydrogenase (LDH) (U/L) | LDH laboratory test within six hours of presentation | N | 155 | 5,000 | 367 | 270-684 | - | 925 | 572-1496 | - |
| - | ANC/ALC ratio | A ratio computed from ANC and ALC | N | 0.027 | 121.3 | 7.04 | 3.99-13.4 | - | 2.77 | 1.4-3.94 | - |

IQR = Interquartile range, N = Numerical, B = Binary, AMS = altered mental status, \*Sex was only needed to categorize lab values

**Figure 2:**
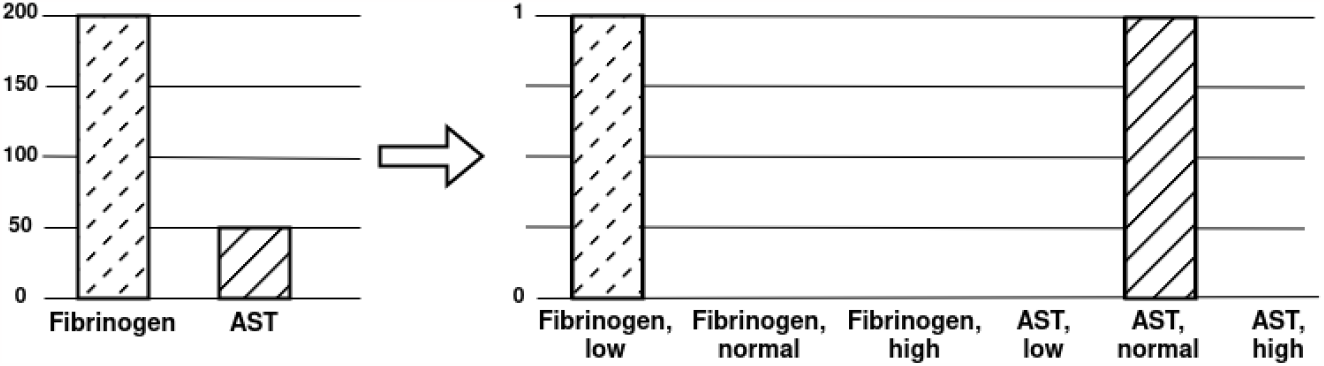
A feature is converted to categorical inputs by using set thresholds

### 3.2 Missing Values

Missing values in datasets is a recurring problem in the healthcare area due to systematic (different data capture protocols used at other times) or non-deterministic (individuals or institutions not providing data) differences [32]. The categorization process based on established thresholds allows handling these missing laboratory values by turning them into zero-value inputs (Fig. 3). These features were converted into a vector with only zeros ([0, 0, 0]), describing the clinical features as neither high, nor normal, nor low, which enabled them to be handled by the deep learning model without the need to use any data imputation method for laboratory features that could create a bias during the learning process.

**Figure 3:**
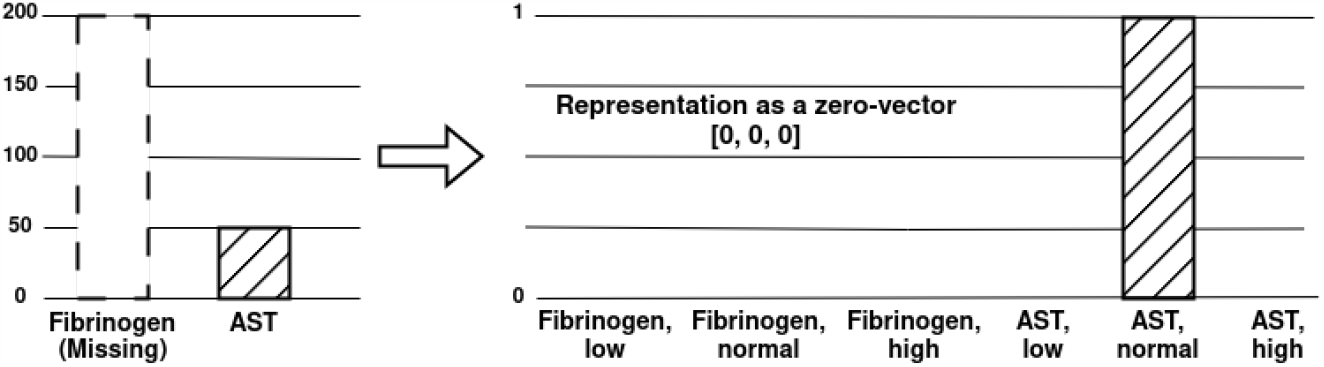
Missing laboratory values are converted into a zero-value vector, enabling them to be handled by the deep learning model.

### 3.3 Contextual Information

The different possible combinations of inputs generated from a feature (low, normal, high, or missing) provided contextual information from patient data generated by converting laboratory features into categorical inputs. That is, the significance of the patient input and its meaning depended not only on its value but on the value of other directly related inputs (inputs generated from the same laboratory feature) (Fig. 4). This design decision was essential when selecting the deep learning model architecture.

**Figure 4:**
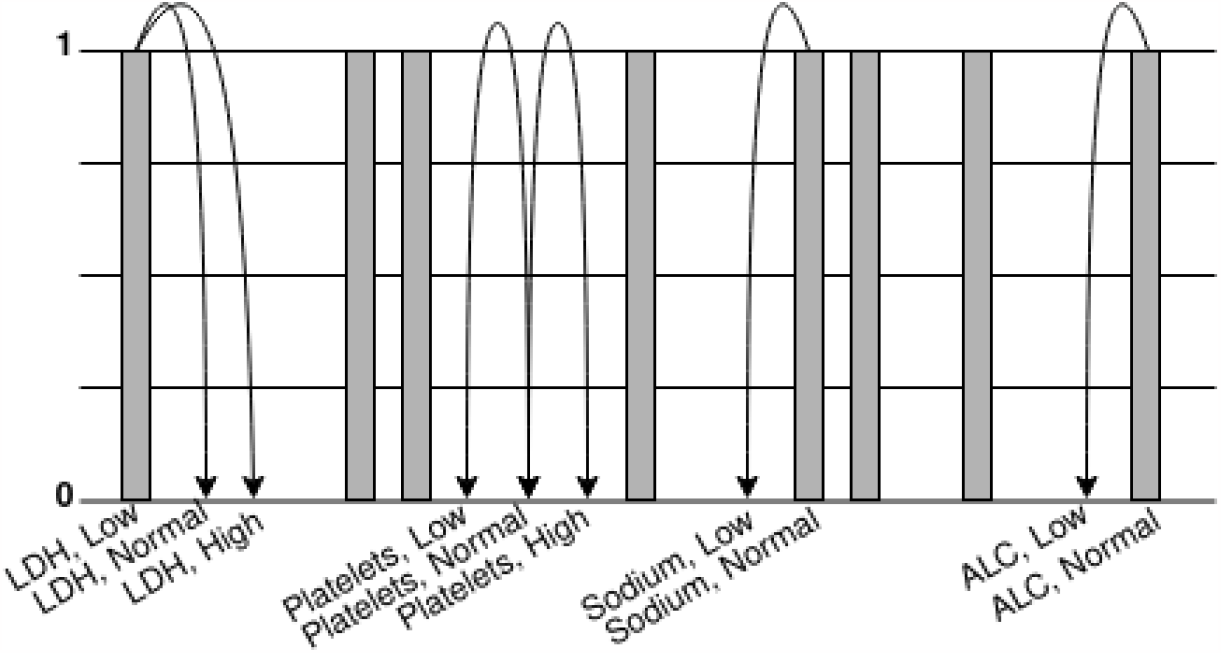
After the categorization process, the inputs are related through the features that they belong to, generating contextual information.

### 3.4 Feature Selection

The 40 out of 73 inputs used were selected through an iterative process based on the values obtained by the attention module of our deep learning model (MET-30). During the model training, the inputs with the lowest values obtained from the attention module were eliminated one by one, while ensuring that the testing accuracy of the model was not reduced. Once the testing accuracy was affected by the elimination of the input with the lowest value obtained, the iterative process of input selection was stopped, resulting in 40 inputs related to 30 features.

### 3.5 MET-17

To create MET-17, we took the attention values of the inputs obtained from the MET-30 attention module and grouped them into four groups using k-means clustering. For all categorical inputs, an attention value *G*_*m*_ was established for each group by taking the arithmetic mean of the attention values of all the inputs in the group. For each categorical input 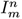 where *m* is the group to which that input belongs and *n* is the input index for that group, the group attention value was multiplied by the difference in the probability of that patient being classified as typhus given that input 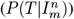 minus the probability of that patient of being classified as MIS-C given the same input 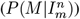 The values obtained were normalized, multiplied by ten, and rounded, obtaining the final scoring values for the categorical inputs as shown in Eq. (1). All values below one were excluded.

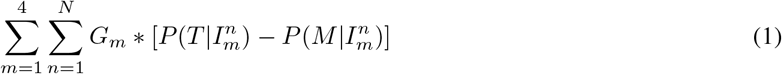

In the case of the continuous values, we used Eq. (2), where a sign function is defined as:

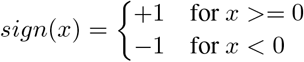

was dependent on an empirically established threshold, 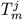 with *m* denoting the attention group and *j* the input in that group. This threshold was set based on the distribution of the value of the input in question for patients with typhus and patients with MIS-C, and its result was multiplied by a constant:

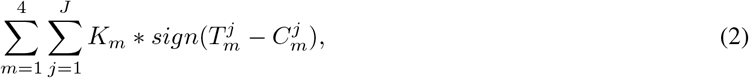

The constant, *K*_*m*_, was determined by the hierarchy shown in Eq. (3), based on which attention value group the input belonged to.

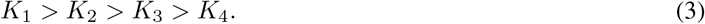

Therefore, the final result for MET-17 consists of the sum of the scores obtained from the categorical Eq. (1) and continuous inputs Eq. (2).

### 3.6 MET-30

In contrast, MET-30 is a Deep Learning model consisting of three components: an attention module capable of providing values that weigh the inputs based on relevance, a long short-term memory (LSTM) layer that can learn the contextual information among the data, and a module of classification composed of dense layers (Fig. 5).

**Figure 5:**
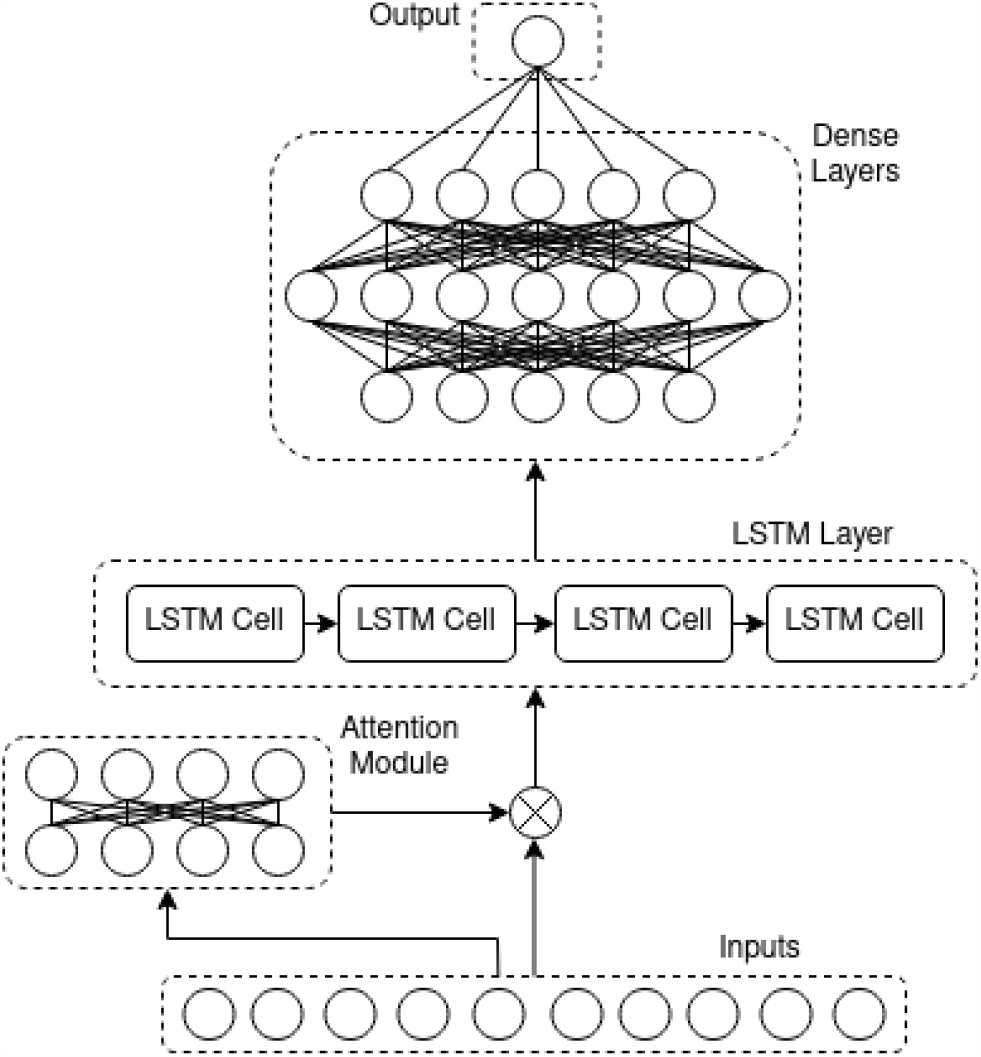
Visualization of the attention LSTM architecture used as part of MET-30.

The attention module of the model weighs the input values, *V*_*n*_, by their importance for the correct classification of patients. This is done by a dense layer, *u*_*n*_, that uses *tanh* as its activation function to maximize the weights of the inputs that contribute the most and minimize the weights of those with the smallest contribution. The next layer, *q*_*n*_, computes the attention values among the inputs using a softmax function, the result of which is multiplied by the inputs to get *r*_*n*_, the LSTM layer inputs:

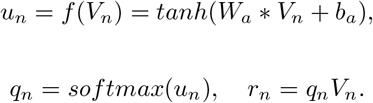

The MET-30 LSTM layer is defined by *i*_*t*_, *f*_*t*_, and *o*_*t*_ that represent the input, forget, and output gates, while *c*_*t*_ is the cell state and *h*_*t*_ the hidden state as defined below:

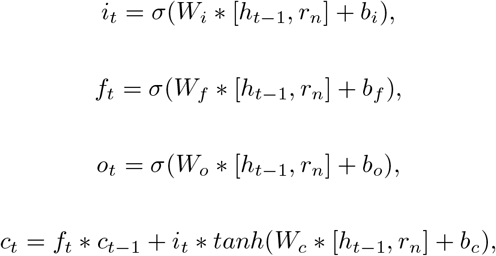

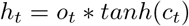

Lastly, the classification module is a set of densely connected layers where every neuron on them is defined by:

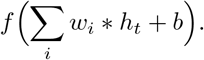

### 3.7 Overfitting

Architectures containing LSTM layers are prone to overfitting [33]. This phenomenon occurs when a machine learning or deep learning model overfits its parameters during its training to “learn” in detail the features present in the training dataset, thus losing the ability to generalize the information and performing poorly on “unseen” data (Fig. 6). To avoid this, a recurrent dropout was implemented in the LSTM layer with a feed-forward dropout in subsequent dense layers of the architecture.

**Figure 6:**
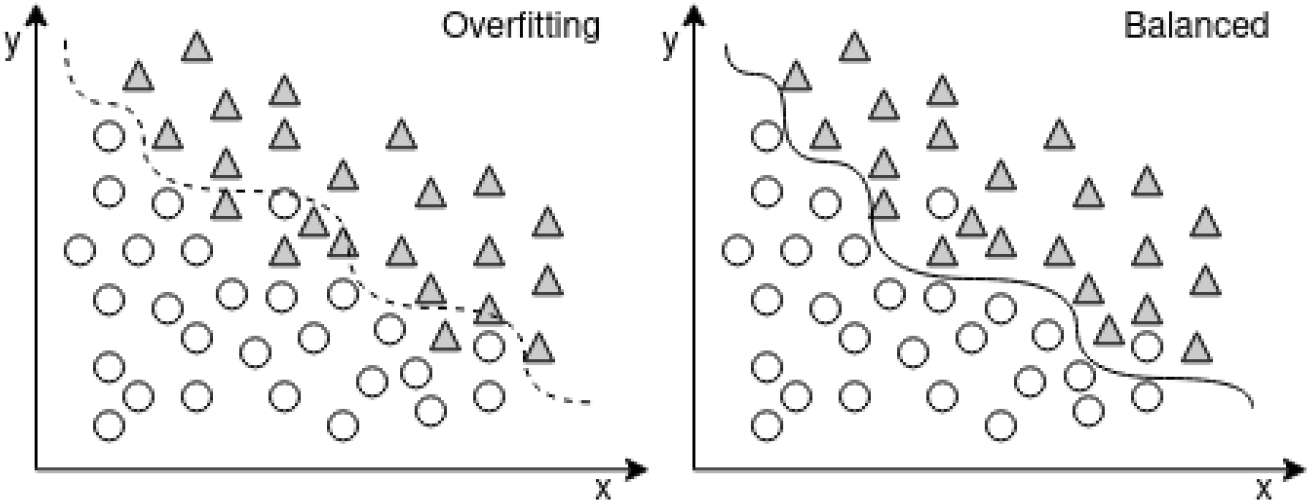
Overfitting diminishes the capability of the model to generalize the data by memorizing the training data.

### 3.8 AI-MET: A Two-stage Decision Support System

AI-MET is designed as a two-stage decision approach. In the first stage, patient information regarding the 17 features used by MET-17 is entered to classify the patient as typhus or MIS-C. If the score obtained, *τ*, is insufficient to issue a reliable diagnosis, 13 more features are requested to be used in MET-30, the deep learning model. To quantify the confidence of the score obtained by MET-17, we designed a Confidence Index (CI) using Eq. (4) for patients with typhus and Eq. (5) for patients with MIS-C. These sigmoid functions were fitted using the parameters *α*, as the bias, *β*, as the coefficient’s maximum value, *δ*, as the function’s growth rate, and *γ*, as the cross-point between both functions, for the distribution of the values obtained from MET-17:

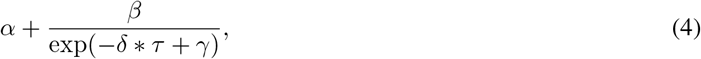

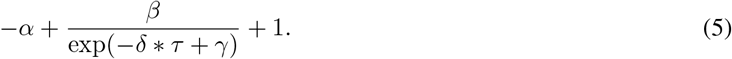

## 4 Results

### 4.1 Training / Testing Dataset Description

The dataset used for training and testing all models included 56 patients admitted with the first surge of MIS-C (May 15 to November 30, 2020), 77 patients admitted with the delta surge of MIS-C (September 1 to October 31, 2021), and 87 patients admitted with murine typhus (January 1, 2020, to December 31, 2021) to TCH and its two satellite campuses within the greater Houston area. Medical records were reviewed, with 49 demographic, clinical, and laboratory features available within six hours of presentation obtained. Of these, 30 features were selected for this study (see subsection III.D).

### 4.2 Validation Dataset Description

The validation dataset included an additional 111 patients with MIS-C admitted between November 2020 and June 2022. Medical records were reviewed for the 17 demographic, clinical, and laboratory features available within six hours of presentation to classify them using MET-17. 54 of the 111 patients obtained a CI equal to or higher than that established and therefore, were classified only using MET-17. For the remaining 57, the 13 additional features were requested to be classified using MET-30.

### 4.3 MET-17: Fixed Points Scoring System

The attention values computed from MET-30 for every input are shown in Fig. 7. The group attention values for categorical inputs, calculated by the arithmetic mean of the attention value belonging to each group, are *G*_1_ = 0.8, *G*_2_ = 0.6, *G*_3_ = 0.4, and *G*_4_ = 0.1. Following the hierarchy established in Eq. (3), the constants for the continuous inputs were arbitrarily established as *K*_1_ = 8, *K*_2_ = 6, *K*_3_ = 4, and *K*_4_ = 1.

**Figure 7:**
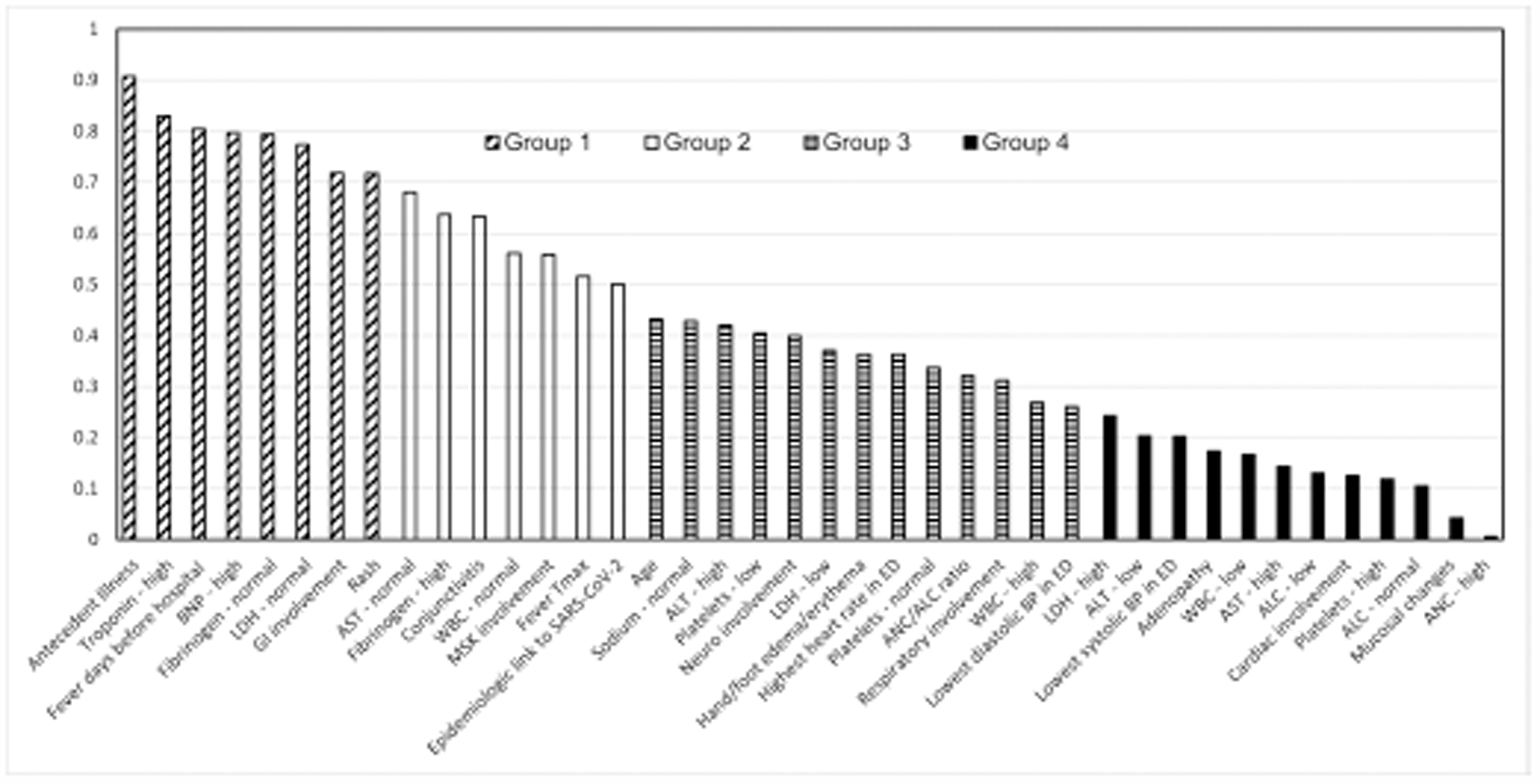
Average scaled attention values obtained from the MET-30 attention module.

The MET-17 fixed point system for laboratory and clinical features are shown in Tables 6 and 7, respectively. The fixed point system for the continuous features can be found in Table 8. Of note, sex was not a weighted feature but was necessary to properly interpret normal laboratory ranges, so it is the 17^*th*^ feature.

**Table 6:** Laboratory features for MET-17

| Feature | If Low | If Normal | If High |
| --- | --- | --- | --- |
| Fibrinogen (mg/dL) | +04 | +16 | -16 |
| Troponin (ng/mL) | +09 | +09 | -09 |
| BNP (pg/mL) | +10 | +10 | -10 |
| ALT (U/L) | -08 | -08 | +08 |
| AST (U/L) | -03 | -11 | +11 |
| Sodium (mmol/L) | -02 | +02 | -02 |

**Table 7:** Clinical features for MET-17

| Feature | If Positive | If Negative |
| --- | --- | --- |
| Epidemiologic link to SARS-CoV-2 | -05 | +05 |
| Rash | +03 | -03 |
| Conjunctivitis | -04 | +04 |
| Antecedent Illness | -02 | +02 |

**Table 8:**
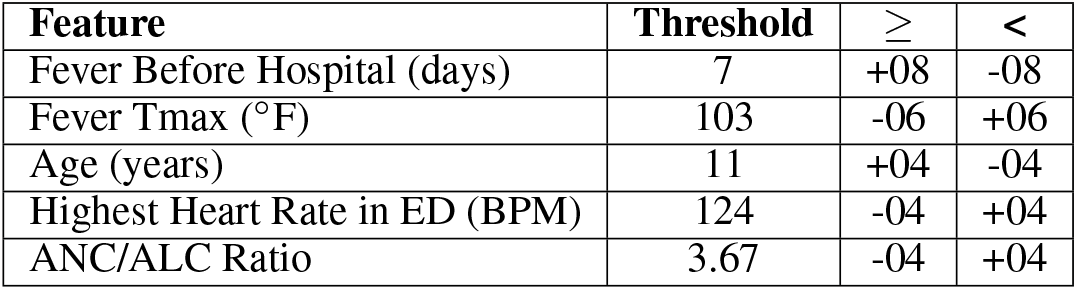
Continuous features for MET-17

### 4.4 Performance Assessment

To evaluate the performance of our approach and the baseline models, we computed True Positive (TP) as a correctly identified typhus patient, True Negative (TN) as a correctly identified MIS-C patient, False Positive (FP) as an MIS-C patient incorrectly identified as a typhus patient, and False Negative (FN) as a typhus patient incorrectly identified as an MIS-C patient. We computed the accuracy, sensitivity, specificity, and precision from these parameters.

### 4.5 Experimental Results

We compared the performance of five statistical and machine learning models: Support Vector Machine (SVM), Dense Neural Network (DNN), Logistic Regression (LR), Random Forrest (RF), and Decision Trees (DT), to our clinical decision support system. The training, testing, and validation of all models were performed with Python 3.9.7, Tensorflow 2.2.0, Pandas 1.1.3, and Keras 2.3.0-tf running on a LINUX-based computer equipped with an AMD Ryzen 5600g CPU and a GeForce RTX3060 GPU. The dataset used for training and testing was evenly divided into cohorts A and B. Information regarding the exact proportion of typhus and MIS-C patients for cohorts A, B, and validation is shown in Table 9. For experiment 1, all models were trained in cohort A and tested in cohort B. For experiment 2, all models were trained in cohort B and tested in cohort A. Results are shown in Fig. 8 and Fig. 9, respectively. Finally, AI-MET was the only model assessed in the validation dataset, obtaining an accuracy of 99 percent.

**Table 9:** Training / Testing and Validation Datasets

| Cohort | Typhus patients | MIS-C patients |
| --- | --- | --- |
| A | 44 | 66 |
| B | 43 | 67 |
| Validation | 0 | 111 |

**Figure 8:**
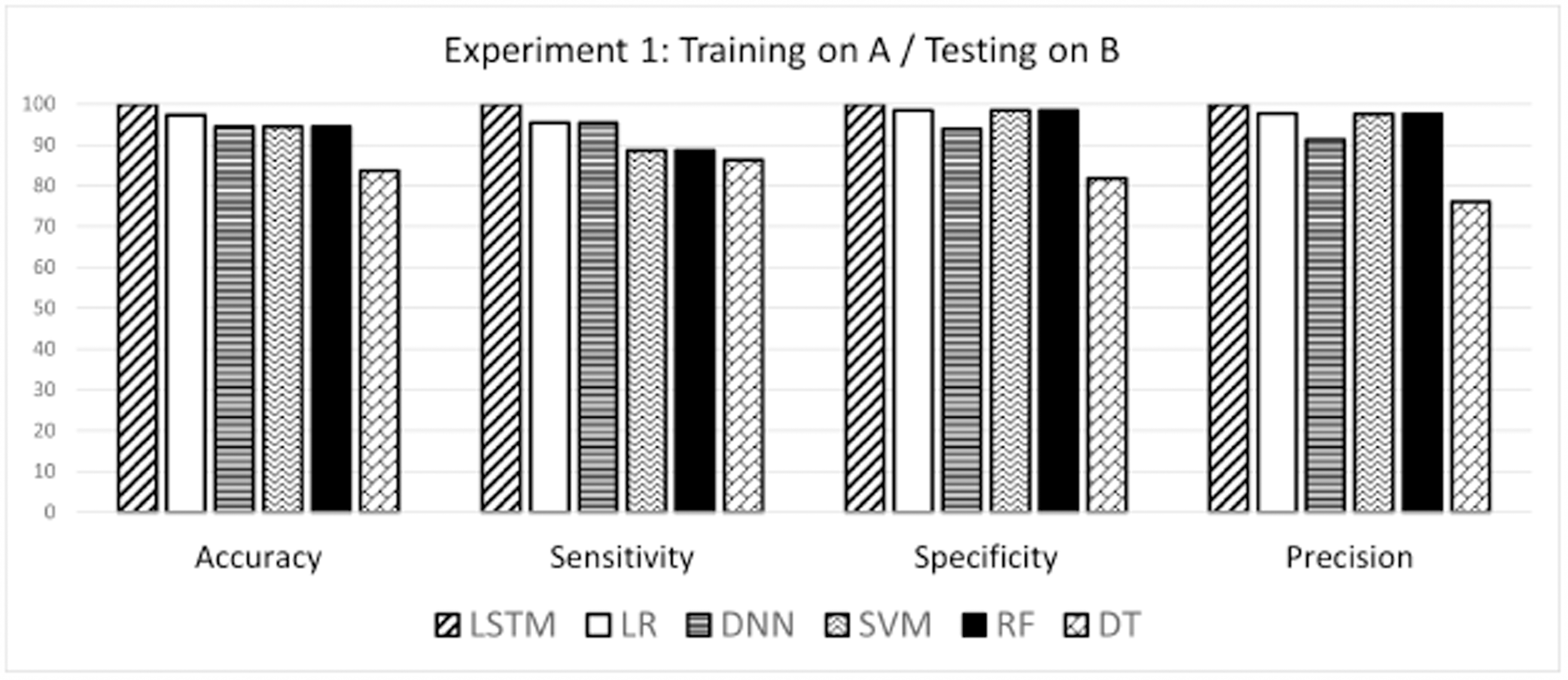
Machine learning models performance for Experiment 1.

**Figure 9:**
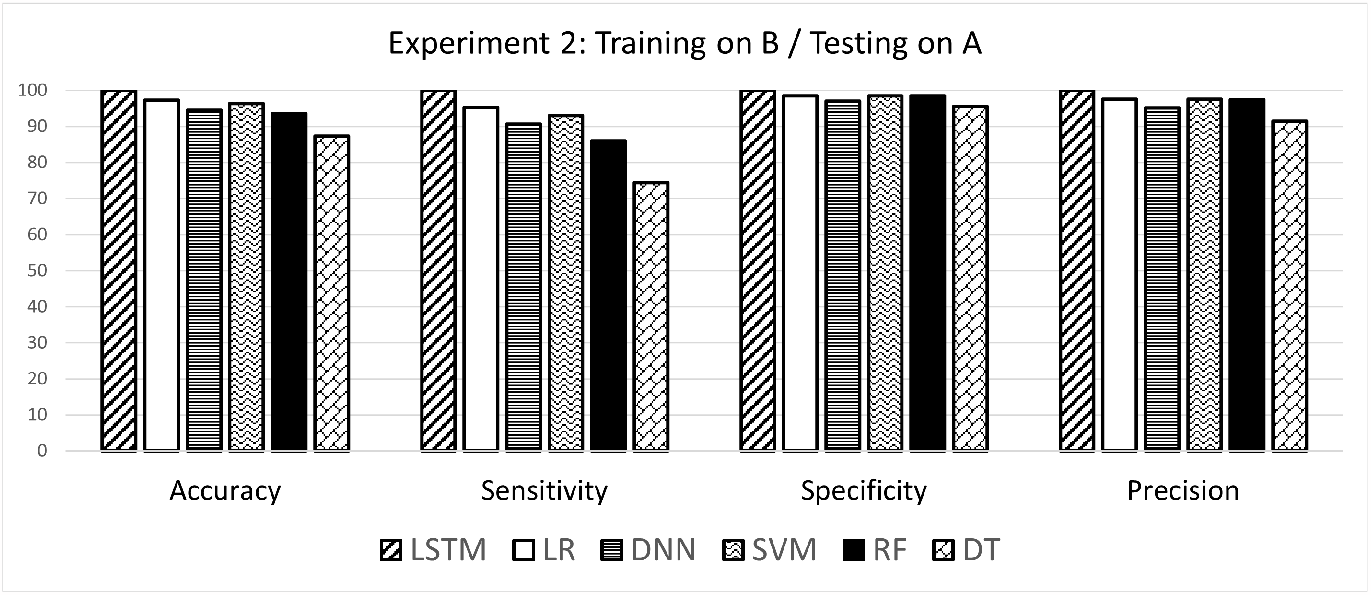
Machine learning models performance for Experiment 2.

## 5 Discussion

Limitations of this work are using a one-center training/testing dataset and only MIS-C patients dataset for validation.

AI-MET outperforms the baseline machine learning models used in our reported metrics. This could be partly due to the integration of the LSTM layer in MET-30, which allows learning of the contextual information generated due to the process of categorizing the laboratory features of the patients.

A well-known drawback of using architectures that contain LSTM layers and the possibility of overfitting is their computational complexity. This can often lead to these architectures being computationally expensive to deploy and requiring higher resource usage. It was for this reason that MET-17 was created. AI-MET provides not only a highly effective deep learning model in the task of distinguishing patients with MIS-C from typhus patients but also a fixed scoring system that can handle approximately half of the patients (Fig. 10) by using the confidence index set at 0.95 in our experiments. In this way, AI-MET becomes a CDSS capable of being implemented in the E.D. without sacrificing accuracy, sensitivity, specificity, or precision.

**Figure 10:**
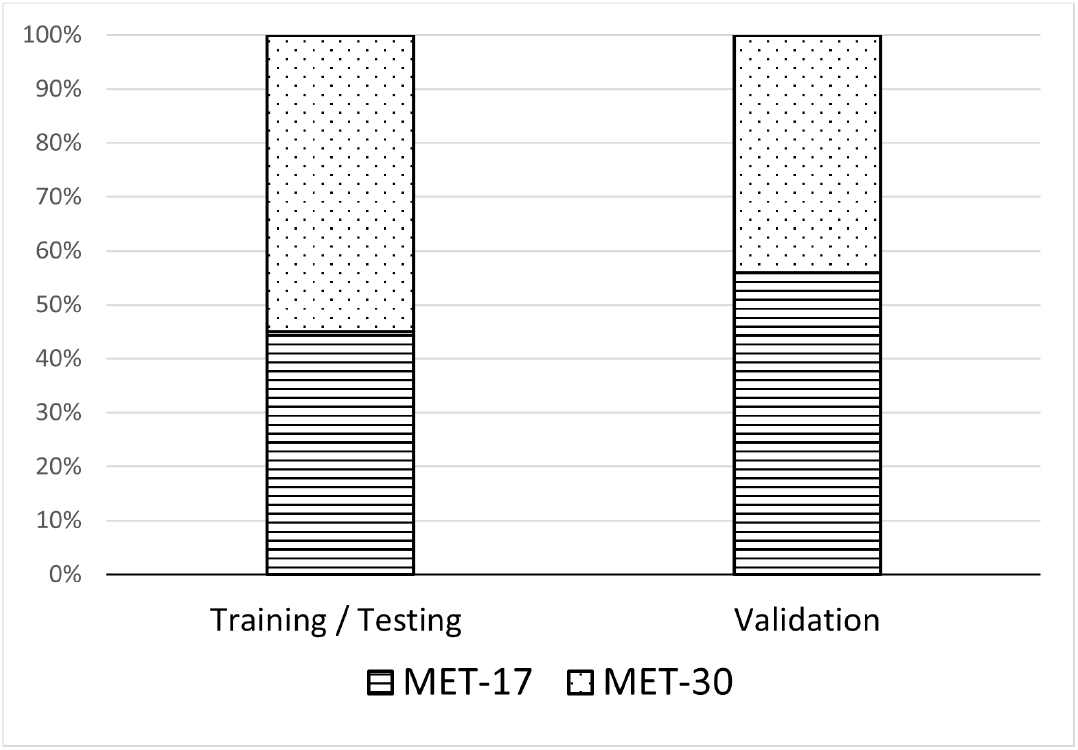
Percentage of patients from each dataset who were classified in each AI-MET stage.

## 6 Conclusion

AI-MET can be successfully employed to distinguish MIS-C from typhus based on features available within the first six hours of patient presentation. Our clinical decision support system optimizes the number of clinical features needed to differentiate MIS-C from typhus with high sensitivity and specificity, and it can be implemented in the Emergency Department with no need for an electronic device in approximately half of the cases.

## Data Availability

All data produced in the present study are available upon reasonable request to the authors.

## Acknowledgment

This work was partly supported by NIH grant R33HD105593. Abraham Bautista-Castillo is also supported by the National Council of Science and Technology of Mexico, scholarship number 739528. Icons and diagrams shown in this work were obtained and created through free licenses from Icons8 [34] and Draw.io [35], respectively. Any opinions, findings, conclusions, or recommendations expressed in this material are those of the authors. They do not necessarily reflect the views of the NIH, other funders, the position, or the policy of the Government, and no official endorsement should be inferred.

